## Supplementary Files for "Developing better digital health measures of Parkinson’s disease using free living data and a crowdsourced data analysis challenge"

**Supplementary Table 1**: P-value for association of top 10 features with label for on/off, dyskinesia and tremor

| | on_off* | dyskinesia* | tremor* |
|---|---|---|---|
| **quantile__q_0.7** | 0.341 | 0.76241 | 0.260796 |
| **number_peaks__n_1** | 0.220 | 0.308912 | 0.453108 |
| **quantile__q_0.2** | 0.4014 | 0.054511 | 0.80456 |
| **quantile__q_0.4** | 0.574647 | 0.757697 | 0.613633 |
| **quantile__q_0.8** | 0.830643 | 0.956247 | 0.872984 |
| **sum_values** | 0.825521 | 0.853557 | 0.677784 |
| **quantile__q_0.3** | 0.850645 | 0.167818 | 0.536281 |
| **fft_coefficient__coeff_0__attr_"real"** | 0.884536 | 0.738591 | 0.89858 |
| **mean** | 0.748474 | 0.919004 | 0.636074 |
| **fft_coefficient__coeff_0__attr_"abs"** | 0.942655 | 0.687168 | 0.575989 |

* Shown are the p-values for testing for association between magnitude of feature SHAP value and label. For each column, a t-test was performed to compare models trained on the column label against models trained on other labels. No tests were significant at $\alpha = 0.05$ implying that none of the features were specific to individual phenotypes.

**Supplementary Table 2**: Association (Kendall's tau) of subject characteristics with model improvement (on/off medication)

| | | dbmi | | HaProzdor | | ROC BEAT-PD | | hecky | | Yuanfang Guan | | Problem Solver | | Meta-Analysis p-val |
|---|---|---|---|---|---|---|---|---|---|---|---|---|---|---|
| | | tau | p-val | tau | p-val | tau | p-val | tau | p-val | tau | p-val | tau | p-val | |
| | n | 0.225 | 0.149 | 0.107 | 0.495 | 0.330 | 0.034 | 0.172 | 0.270 | 0.234 | 0.134 | 0.189 | 0.224 | 0.158 |
| | Age | 0.048 | 0.804 | 0.048 | 0.804 | -0.106 | 0.585 | 0.144 | 0.457 | -0.144 | 0.457 | 0.144 | 0.457 | 0.905 |
| UPDRS | Part I | 0.250 | 0.219 | 0.333 | 0.102 | 0.083 | 0.682 | 0.270 | 0.183 | 0.250 | 0.219 | 0.499 | 0.014 | 0.131 |
| | Part II | 0.217 | 0.272 | 0.276 | 0.162 | 0.059 | 0.764 | 0.177 | 0.369 | 0.039 | 0.842 | 0.177 | 0.369 | 0.399 |
| | Part IV | 0.000 | 1.000 | -0.039 | 0.842 | 0.020 | 0.920 | -0.099 | 0.617 | 0.158 | 0.424 | 0.375 | 0.058 | 0.710 |
| | Part III (Off) | 0.260 | 0.221 | 0.416 | 0.050 | 0.286 | 0.178 | 0.312 | 0.142 | 0.338 | 0.111 | 0.260 | 0.221 | 0.127 |
| | Part III (On) | 0.107 | 0.621 | 0.267 | 0.217 | 0.080 | 0.711 | 0.107 | 0.621 | 0.053 | 0.805 | -0.107 | 0.621 | 0.680 |
| | Reporting Lag | -0.067 | 0.770 | -0.105 | 0.626 | 0.067 | 0.770 | -0.048 | 0.846 | 0.124 | 0.559 | 0.105 | 0.626 | 0.946 |
| | Label Variance | 0.543 | 0.004 | 0.429 | 0.027 | 0.486 | 0.011 | 0.448 | 0.021 | 0.543 | 0.004 | 0.410 | 0.036 | 0.010 |

**Supplementary Table 3**: Association (Kendall's tau) of subject characteristics with model improvement (dyskinesia)

| | | ROC BEAT-PD | | Yuanfang Guan | | hecky | | Meta-Analysis p-val |
|---|---|---|---|---|---|---|---|---|
| | | **tau** | **p-val** | **tau** | **p-val** | **tau** | **p-val** | |
| | n | -0.202 | 0.279 | 0.034 | 0.857 | -0.134 | 0.470 | 0.692 |
| | Age | -0.537 | 0.023 | -0.278 | 0.240 | -0.241 | 0.309 | 0.273 |
| UPDRS | Part I | -0.101 | 0.681 | 0.060 | 0.805 | -0.141 | 0.565 | 0.856 |
| | Part II | -0.305 | 0.205 | -0.229 | 0.342 | -0.076 | 0.751 | 0.529 |
| | Part IV | -0.057 | 0.813 | -0.094 | 0.694 | -0.623 | 0.009 | 0.417 |
| | Part III (Off) | 0.386 | 0.125 | 0.114 | 0.652 | 0.205 | 0.417 | 0.496 |
| | Part III (On) | 0.092 | 0.717 | 0.138 | 0.587 | 0.414 | 0.103 | 0.533 |
| | Reporting Lag | -0.309 | 0.218 | -0.418 | 0.087 | -0.455 | 0.060 | 0.220 |
| | Label Variance | 0.636 | 0.006 | 0.382 | 0.121 | 0.127 | 0.648 | 0.231 |

**Supplementary Table 4**: Association (Kendall's tau) of subject characteristics with model improvement (tremor)

| | | Yuanfang Guan | | dbmi | | ROC BEAT-PD | | HaProzdor | | Problem Solver | | hecky | | Meta-Analysis p-val |
|---|---|---|---|---|---|---|---|---|---|---|---|---|---|---|
| | | **tau** | **p-val** | **tau** | **p-val** | **tau** | **p-val** | **tau** | **p-val** | **tau** | **p-val** | **tau** | **p-val** | |
| | n | -0.131 | 0.440 | -0.190 | 0.261 | 0.083 | 0.623 | 0.000 | 1.000 | 0.202 | 0.233 | -0.024 | 0.888 | 0.951 |
| | Age | -0.194 | 0.359 | 0.116 | 0.582 | -0.090 | 0.669 | -0.260 | 0.221 | -0.297 | 0.160 | -0.271 | 0.199 | 0.417 |
| UPDRS | Part I | 0.406 | 0.062 | 0.352 | 0.106 | 0.406 | 0.062 | 0.123 | 0.575 | 0.433 | 0.046 | 0.379 | 0.081 | 0.086 |
| | Part II | 0.278 | 0.196 | 0.464 | 0.031 | 0.066 | 0.758 | 0.133 | 0.537 | 0.066 | 0.758 | 0.013 | 0.951 | 0.404 |
| | Part IV | 0.225 | 0.295 | -0.066 | 0.758 | 0.331 | 0.123 | -0.187 | 0.387 | 0.331 | 0.123 | -0.013 | 0.951 | 0.611 |
| | Part III (Off) | 0.330 | 0.160 | 0.257 | 0.274 | 0.183 | 0.435 | 0.537 | 0.023 | 0.294 | 0.212 | 0.330 | 0.160 | 0.157 |
| | Part III (On) | -0.075 | 0.753 | 0.112 | 0.637 | 0.000 | 1.000 | 0.321 | 0.180 | -0.112 | 0.637 | 0.037 | 0.875 | 0.836 |
| | Reporting Lag | 0.179 | 0.435 | 0.179 | 0.435 | 0.385 | 0.076 | -0.039 | 0.855 | 0.282 | 0.204 | -0.154 | 0.510 | 0.699 |
| | Label Variance | 0.590 | 0.004 | 0.282 | 0.204 | 0.282 | 0.204 | 0.090 | 0.669 | 0.385 | 0.076 | 0.205 | 0.367 | 0.132 |

### Supplementary Table 5: Validation of models in clinically labeled segments (on/off medication)

| | dbmi | | haProzdor | | ROC BEAT-PD | | Yuanfang Guan | |
|---|---|---|---|---|---|---|---|---|
| **Subject ID** | **Correlation** | **P-value*** | **Correlation** | **P-value** | **Correlation** | **P-value** | **Correlation** | **P-value** |
| 1004 | 0.324 | 0.002 | 0.387 | 1.41E-04 | 0.316 | 0.002 | 0.493 | 4.76E-06 |
| 1007 | 0.176 | 0.111 | 0.109 | 0.212 | 0.046 | 0.369 | -0.093 | 0.742 |
| 1019 | 0.036 | 0.365 | -0.015 | 0.557 | 0.335 | 3.69E-04 | -0.166 | 0.943 |
| 1020 | -0.006 | 0.508 | 0.271 | 0.174 | 0.296 | 0.152 | -0.028 | 0.537 |
| 1023 | -0.051 | 0.689 | -0.063 | 0.733 | 0.033 | 0.372 | 0.207 | 0.021 |
| 1032 | 0.348 | 2.59E-04 | -0.040 | 0.650 | -0.292 | 0.998 | 0.040 | 0.349 |
| 1038 | 0.146 | 0.092 | 0.225 | 0.019 | -0.187 | 0.957 | 0.495 | 9.80E-07 |
| 1039 | 0.275 | 0.004 | -0.002 | 0.507 | 0.045 | 0.331 | 0.027 | 0.397 |
| 1043 | 0.377 | 6.58E-05 | 0.268 | 0.004 | 0.144 | 0.079 | 0.336 | 4.96-04 |
| 1044 | 0.361 | 1.64E-04 | NA** | NA | -0.188 | 0.968 | 0.092 | 0.188 |
| 1046 | NA | NA | NA | NA | 0.152 | 0.067 | NA | NA |
| 1048 | 0.258 | 0.011 | 0.372 | 3.11E-04 | -0.109 | 0.833 | 0.274 | 0.008 |
| 1049 | -0.192 | 0.960 | -0.032 | 0.612 | -0.186 | 0.955 | 0.017 | 0.439 |
| 1051 | 0.280 | 0.003 | -0.033 | 0.628 | 0.410 | 1.40E-05 | -0.367 | 1.000 |
| **Meta-Analysis*** ** | | **1.80e-12** | | **1.04e-05** | | **9.81e-06** | | **6.28E-10** |

* One-sided *p*-value
** NA indicates that a model could not produce a prediction for this individual or that the prediction was constant for all segments
*** Unadjusted *p*-value

**Supplementary Table 6: Validation of models in clinically labeled segments (dyskinesia)**

| | ROC BEAT-PD | | Yuanfang Guan | | HaProzdor | | dbmi | |
|---|---|---|---|---|---|---|---|---|
| **Subject ID** | **Correlation** | **P-value*** | **Correlation** | **P-value** | **Correlation** | **P-value** | **Correlation** | **P-value** |
| 1004 | -0.231 | 0.986 | -0.371 | 1.000 | -0.131 | 0.890 | -0.306 | 0.998 |
| 1020 | -0.299 | 0.861 | NA* | NA | NA | NA | NA | NA |
| 1023 | 0.107 | 0.138 | -0.034 | 0.634 | NA | NA | 0.075 | 0.224 |
| 1039 | 0.115 | 0.121 | 0.015 | 0.438 | 0.047 | 0.318 | -0.011 | 0.541 |
| 1043 | 0.068 | 0.245 | -0.229 | 0.990 | NA | NA | 0.051 | 0.301 |
| 1048 | 0.286 | 0.003 | 0.118 | 0.136 | NA | NA | 0.290 | 0.004 |
| 1049 | 0.049 | 0.321 | -0.076 | 0.762 | -0.155 | 0.929 | -0.004 | 0.513 |
| **Meta-Analysis*** ** | | **0.035** | | **0.850** | | **0.849** | | **0.082** |

* One-sided *p*-values reported
** NA indicates that a model could not produce a prediction for this individual or that the prediction was constant for all segments
*** Unadjusted *p*-value

**Supplementary Table 7: Validation of models in clinically labeled segments (tremor)**

| | Yuanfang Guan | | dbmi | | ROC BEAT-PD | | haProzdor | |
|---|---|---|---|---|---|---|---|---|
| **Subject ID** | **Correlation** | **P-value*** | **Correlation** | **P-value** | **Correlation** | **P-value** | **Correlation** | **P-value** |
| 1004 | 0.145 | 0.086 | 0.215 | 0.024 | 0.032 | 0.384 | 0.480 | 8.38E-07 |
| 1007 | -0.038 | 0.614 | 0.025 | 0.428 | -0.043 | 0.627 | NA | NA |
| 1019 | -0.050 | 0.694 | -0.078 | 0.780 | -0.146 | 0.931 | -0.059 | 0.724 |
| 1020 | 0.159 | 0.286 | 0.002 | 0.497 | 0.168 | 0.275 | 0.171 | 0.271 |
| 1023 | -0.049 | 0.690 | -0.040 | 0.658 | -0.010 | 0.539 | NA | NA |
| 1032 | -0.050 | 0.696 | 0.392 | 1.95E-05 | 0.100 | 0.155 | 0.037 | 0.353 |
| 1038 | 0.081 | 0.222 | 0.178 | 0.045 | 0.040 | 0.352 | 0.018 | 0.432 |
| 1039 | 0.009 | 0.462 | NA | NA | 0.122 | 0.108 | NA | NA |
| 1043 | 0.124 | 0.104 | 0.256 | 0.004 | 0.126 | 0.101 | 0.200 | 0.021 |
| 1046 | 0.267 | 0.003 | -0.305 | 0.999 | 0.077 | 0.216 | NA | NA |
| 1048 | 0.099 | 0.179 | 0.509 | 3.24E-07 | -0.183 | 0.956 | -0.110 | 0.846 |
| 1049 | 0.096 | 0.183 | 0.273 | 0.004 | -0.172 | 0.949 | 0.043 | 0.341 |
| 1051 | -0.027 | 0.606 | NA | NA | 0.093 | 0.173 | NA | NA |
| **Meta-Analysis*** ** | | **0.047** | | **1.97e-10** | | **0.337** | | **1.27e-04** |

* One-sided *p*-value
** NA indicates that a model could not produce a prediction for this individual or that the prediction was constant for all segments
*** Unadjusted *p*-value

**Supplementary Table 8: REAL-PD medication status harmonization**

| Original category | On/off | Dyskinesia |
|---|---|---|
| Off | 1 | 0 |
| On without dyskinesia | 0 | 0 |
| On with non-troublesome dyskinesia | 0 | 1 |
| On with severe dyskinesia | 0 | 2 |

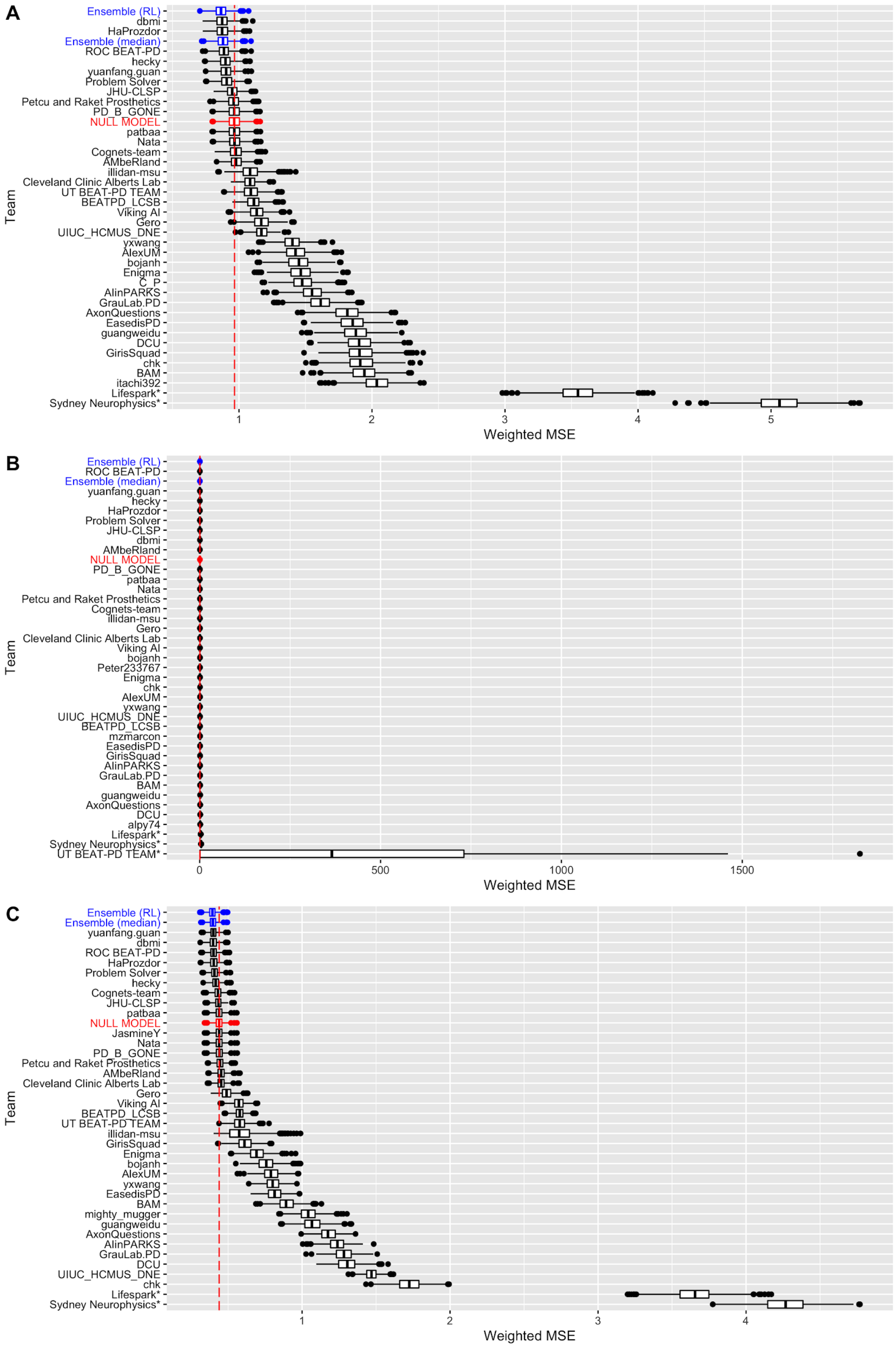
A
Ensemble (RL)
dbmi
HaProzdor
Ensemble (median)
ROC BEAT-PD
hecky
yuanfang.guan
Problem Solver
JHU-CLSP
Petcu and Raket Prosthetics
PD_B_GONE
NULL MODEL
patbaa
Nata
Cognets-team
AMbeRland
illidan-msu
Cleveland Clinic Alberts Lab
UT BEAT-PD TEAM
BEATPD_LCSB
Viking AI
Gero
UIUC_HCMUS_DNE
yxwang
AlexUM
bojanh
Enigma
C_P
AlinPARKS
GrauLab.PD
AxonQuestions
EasedisPD
guangweidu
DCU
GirisSquad
chk
BAM
itachi392
Lifespark*
Sydney Neurophysics*
Team
Weighted MSE
B
Ensemble (RL)
ROC BEAT-PD
Ensemble (median)
yuanfang.guan
hecky
HaProzdor
Problem Solver
JHU-CLSP
dbmi
AMbeRland
NULL MODEL
PD_B_GONE
patbaa
Nata
Petcu and Raket Prosthetics
Cognets-team
illidan-msu
Gero
Cleveland Clinic Alberts Lab
Viking AI
bojanh
Peter233767
Enigma
chk
AlexUM
yxwang
UIUC_HCMUS_DNE
BEATPD_LCSB
mzmarcon
EasedisPD
GirisSquad
AlinPARKS
GrauLab.PD
BAM
guangweidu
AxonQuestions
DCU
alpy74
Lifespark*
Sydney Neurophysics*
UT BEAT-PD TEAM*
Team
Weighted MSE
C
Ensemble (RL)
Ensemble (median)
yuanfang.guan
dbmi
ROC BEAT-PD
HaProzdor
Problem Solver
hecky
Cognets-team
JHU-CLSP
patbaa
NULL MODEL
JasmineY
Nata
PD_B_GONE
Petcu and Raket Prosthetics
AMbeRland
Cleveland Clinic Alberts Lab
Gero
Viking AI
BEATPD_LCSB
UT BEAT-PD TEAM
illidan-msu
GirisSquad
Enigma
bojanh
AlexUM
yxwang
EasedisPD
BAM
mighty_mugger
guangweidu
AxonQuestions
AlinPARKS
GrauLab.PD
DCU
UIUC_HCMUS_DNE
chk
Lifespark*
Sydney Neurophysics*
Team
Weighted MSE

**Supplementary Fig 1**: Bootstraps (n = 1000) of submissions for (A) SC1: on/off, (B) SC2: dyskinesia, and (C) SC3: tremor. Team models (black) and their ensembles (blue) are ordered by rank. Boxes correspond to the $25^{th}$, $50^{th}$, and $75^{th}$ percentiles, and individual points are displayed beyond 1.5*IQR (interquartile range) from the edge of the box.  For each sub-challenge, a null model (show in red) estimated as the subject-specific mean of the training labels was used as a benchmark.

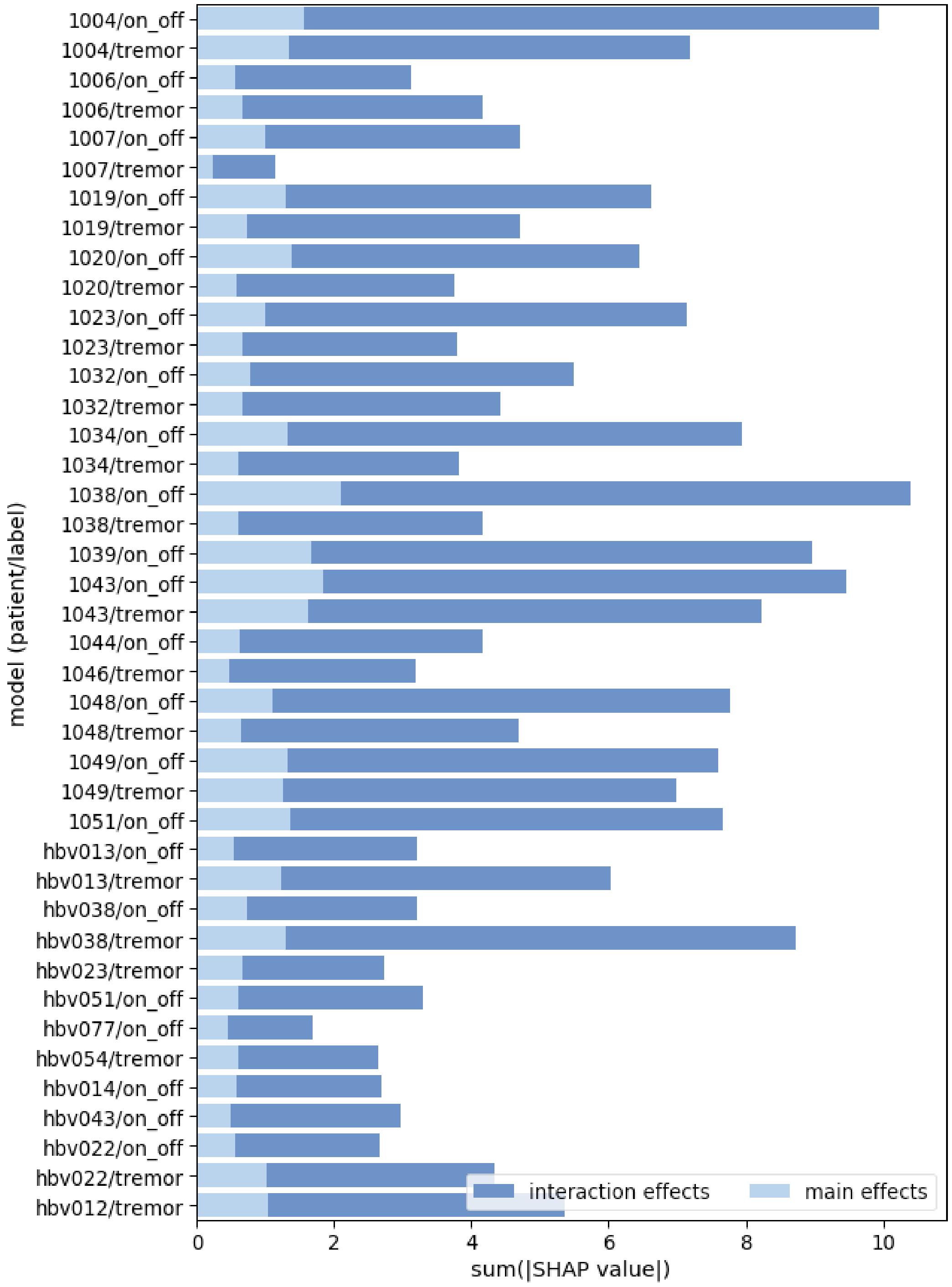


**Supplementary Figure 2**: Sum of magnitudes of SHAP interaction values with main and interaction effects separately shown. Interaction effects outweigh main effects in all models

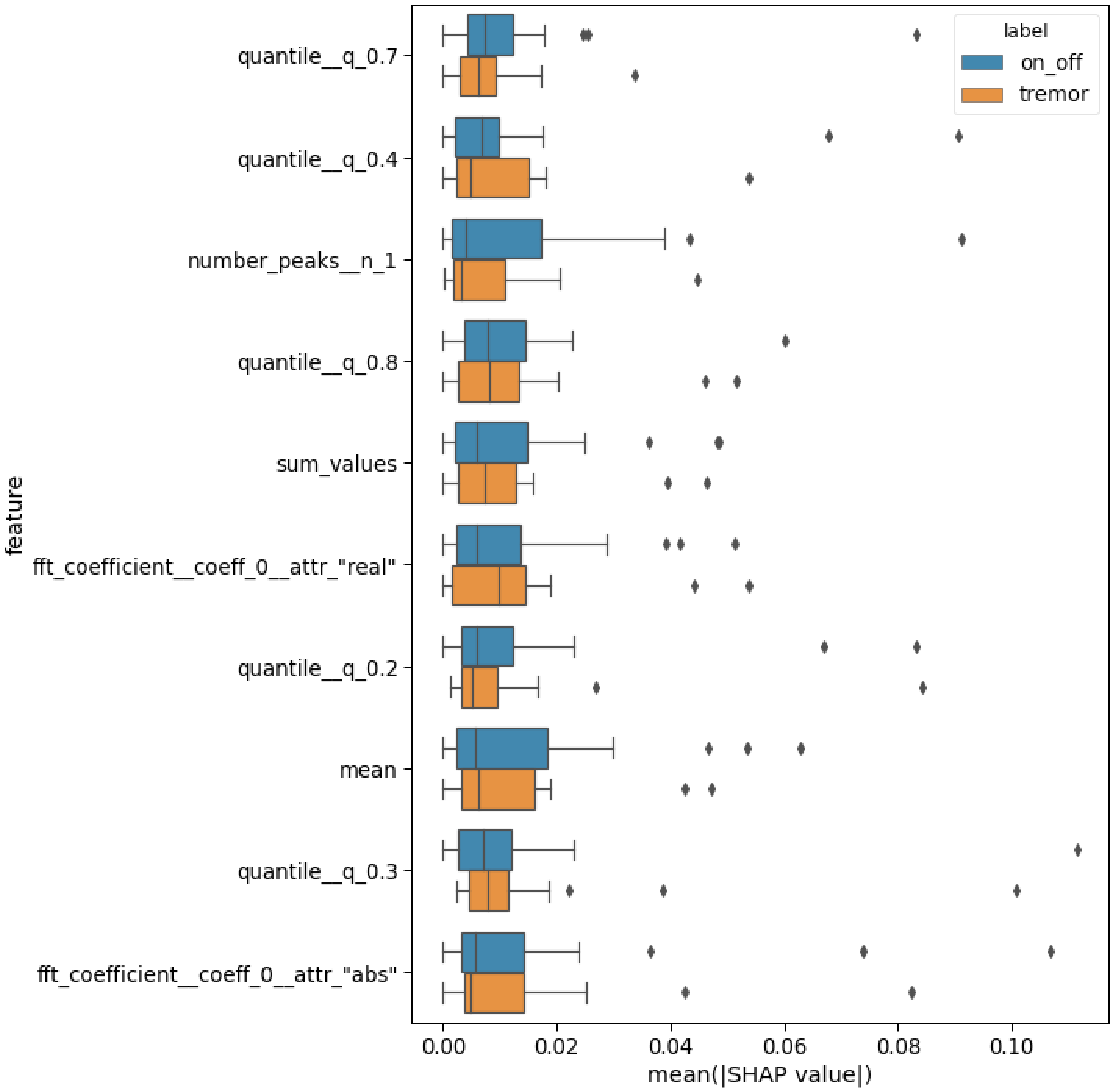


**Supplementary Figure 3:** Distributions of SHAP magnitude for the top 10 features over models for *tremor* and *on_off* labels. *quantile__q_{0.2, 0.3, 0.4, 0.7, 0.8}* are the 20, 30, 40, 70, and 80th data percentiles, respectively; *number_peaks__n_1* is defined as the number of peaks of at least support 1; *sum_values* is the sum over timeseries values; *mean* is the mean timeseries value; and *fft_coefficient__coeff_0__attr_{"real", "abs"}* are the real component and absolute value of the 0th coefficient of the fast Fourier Transform (0 Hz), respectively.

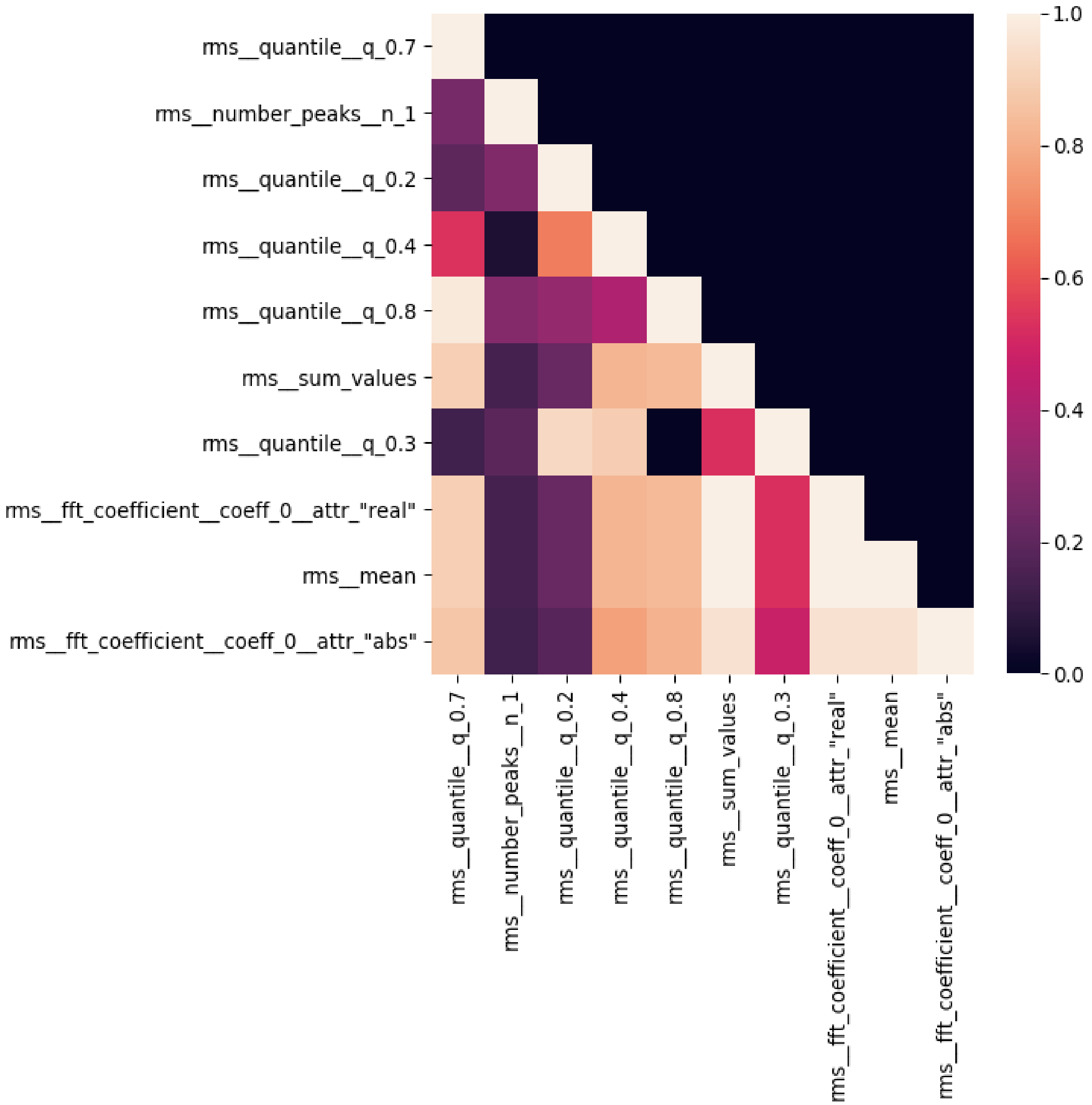


**Supplementary Figure 4:** Heatmap indicating strong correlation among top 10 features. Some features are definitionally equivalent (eg 0 Hz FFT component and mean), while others are very similar in definition (eg 30th and 40th percentiles) contributing to strong correlation.

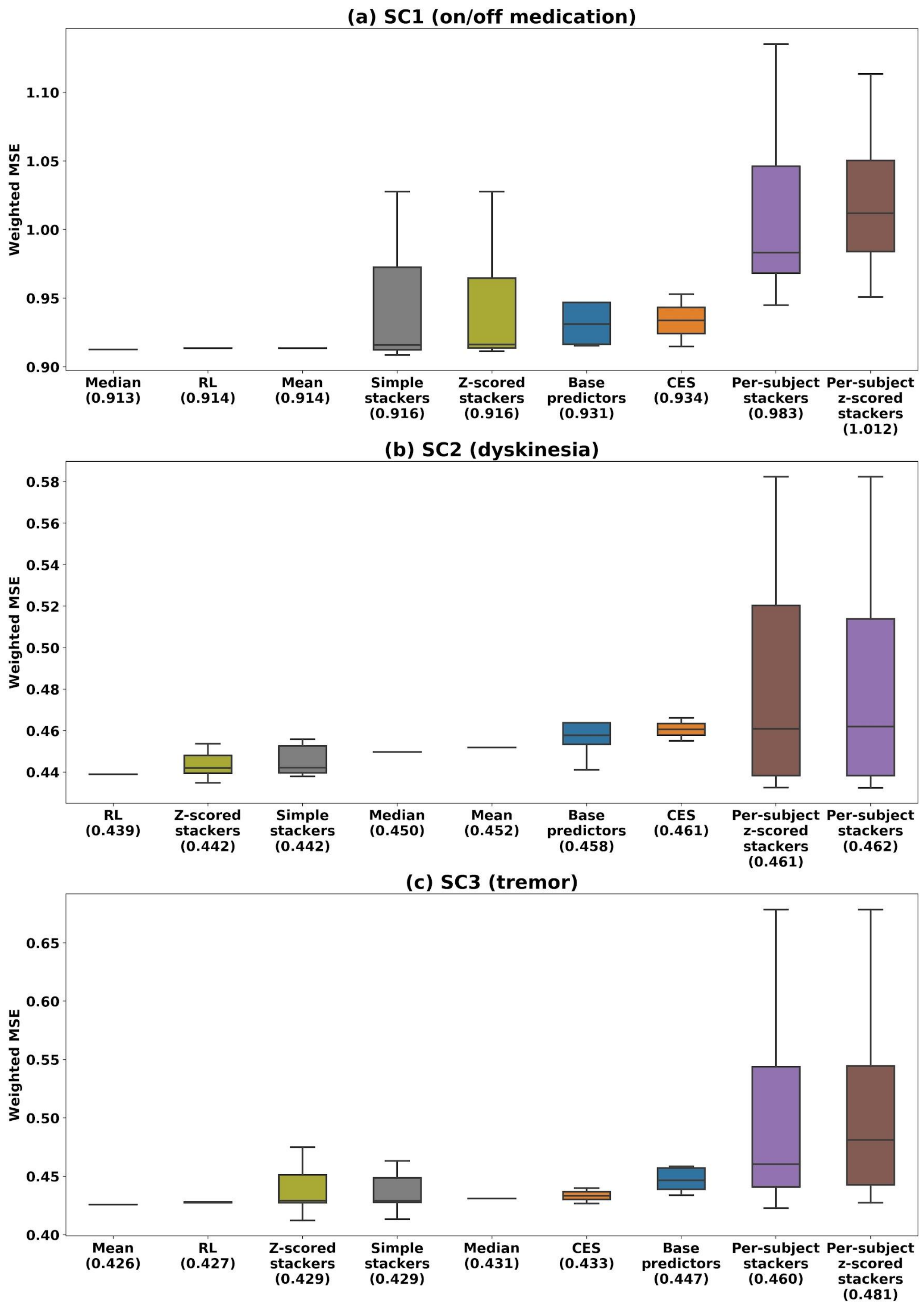


**Supplementary Figure 5**: Distributions of performance of the various categories of ensembles constructed for SC1-3 on the corresponding validation (6$^{th}$) fold of the corresponding training sets. Also shown are the median scores of each category, as well as those of the five base predictors generated by the individual teams' methods.

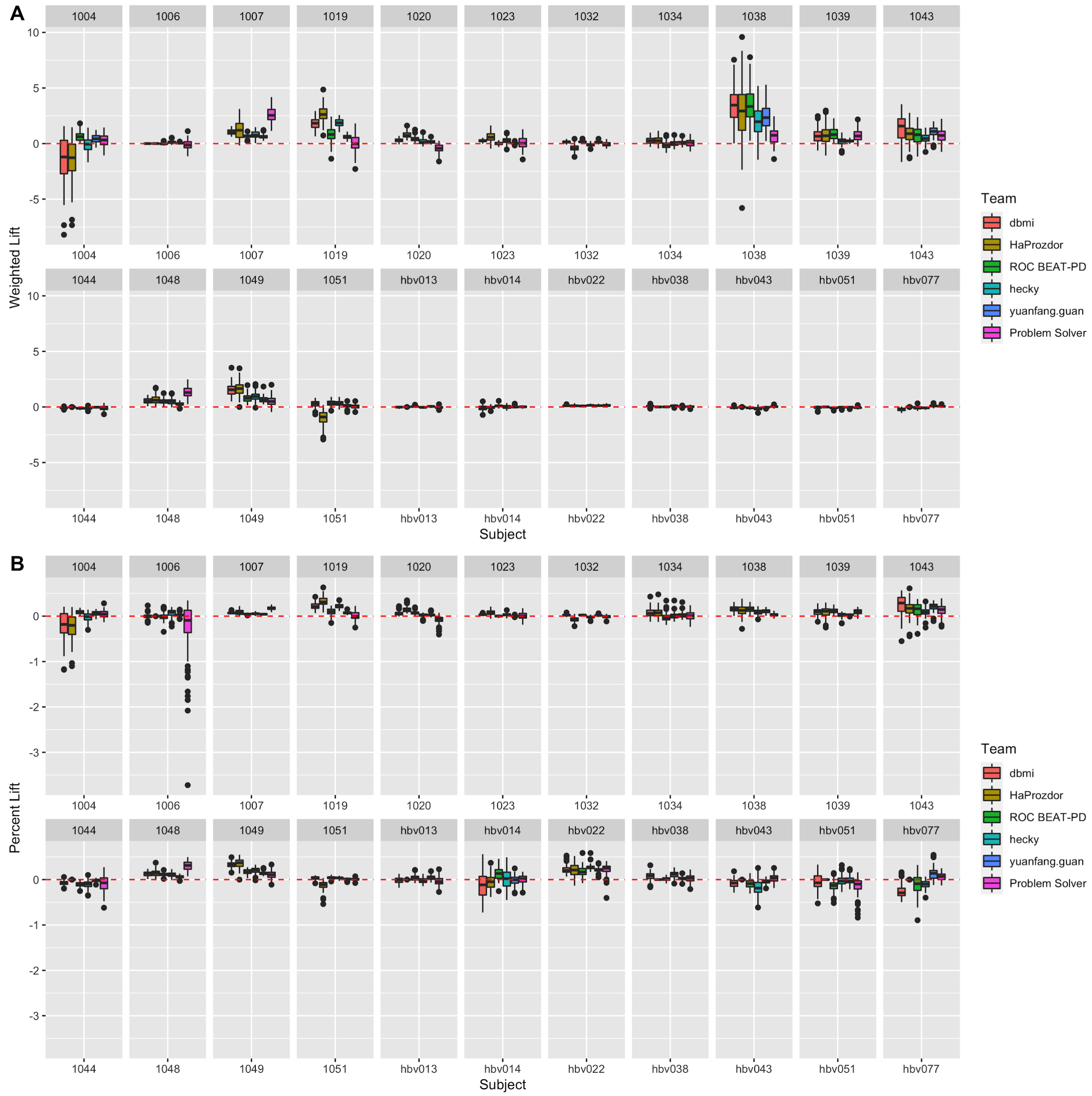


**Supplementary Figure 6:** On/off medication subject-specific lift. (A) Weighted by sqrt(n) and (B) as a percentage of the null model MSE. Teams are ordered by overall rank.

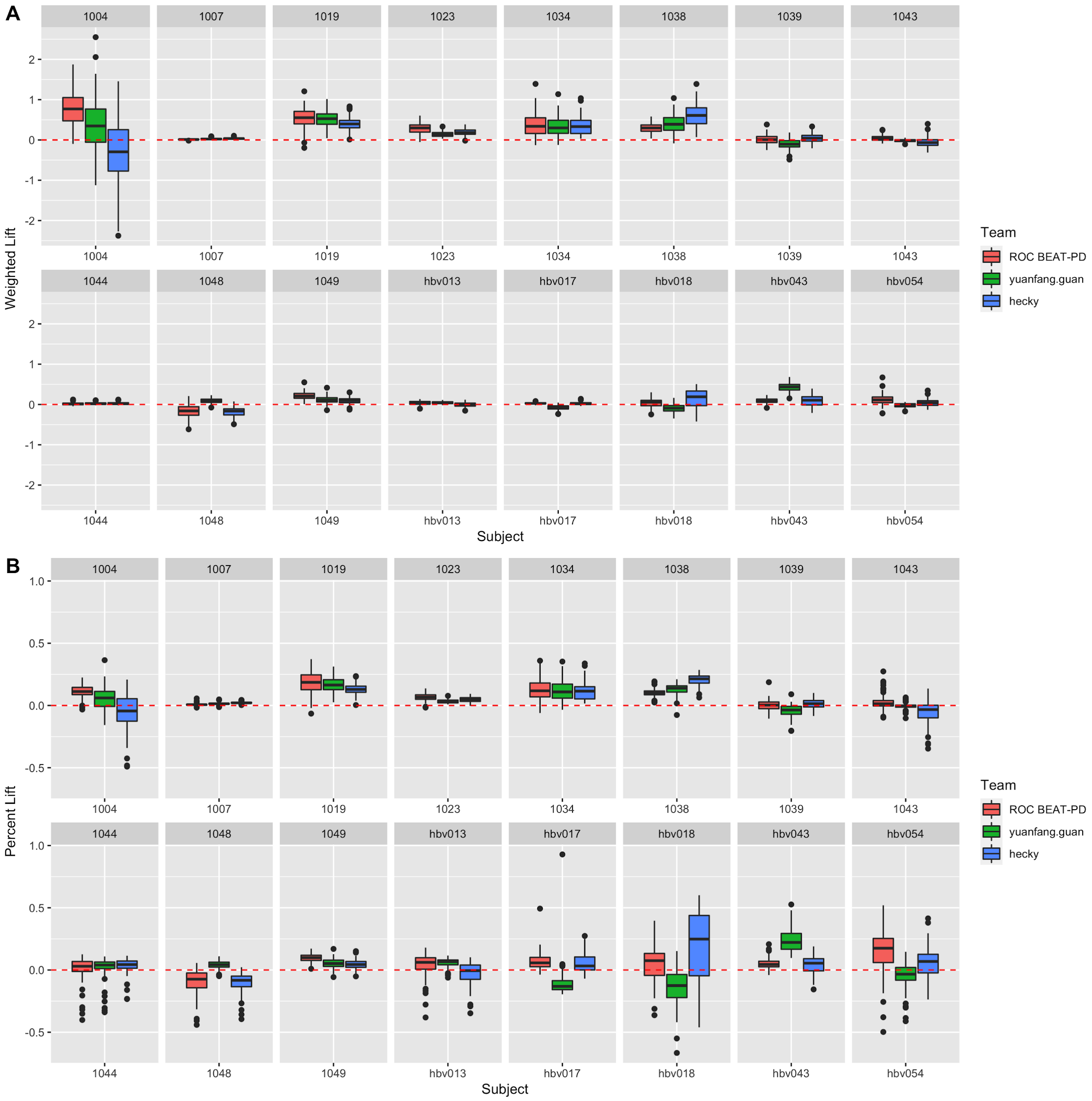


**Supplementary Figure 7**: Dyskinesia subject-specific lift. (A) Weighted by sqrt(n) and (B) as a percentage of the null model MSE. Teams are ordered by overall rank.

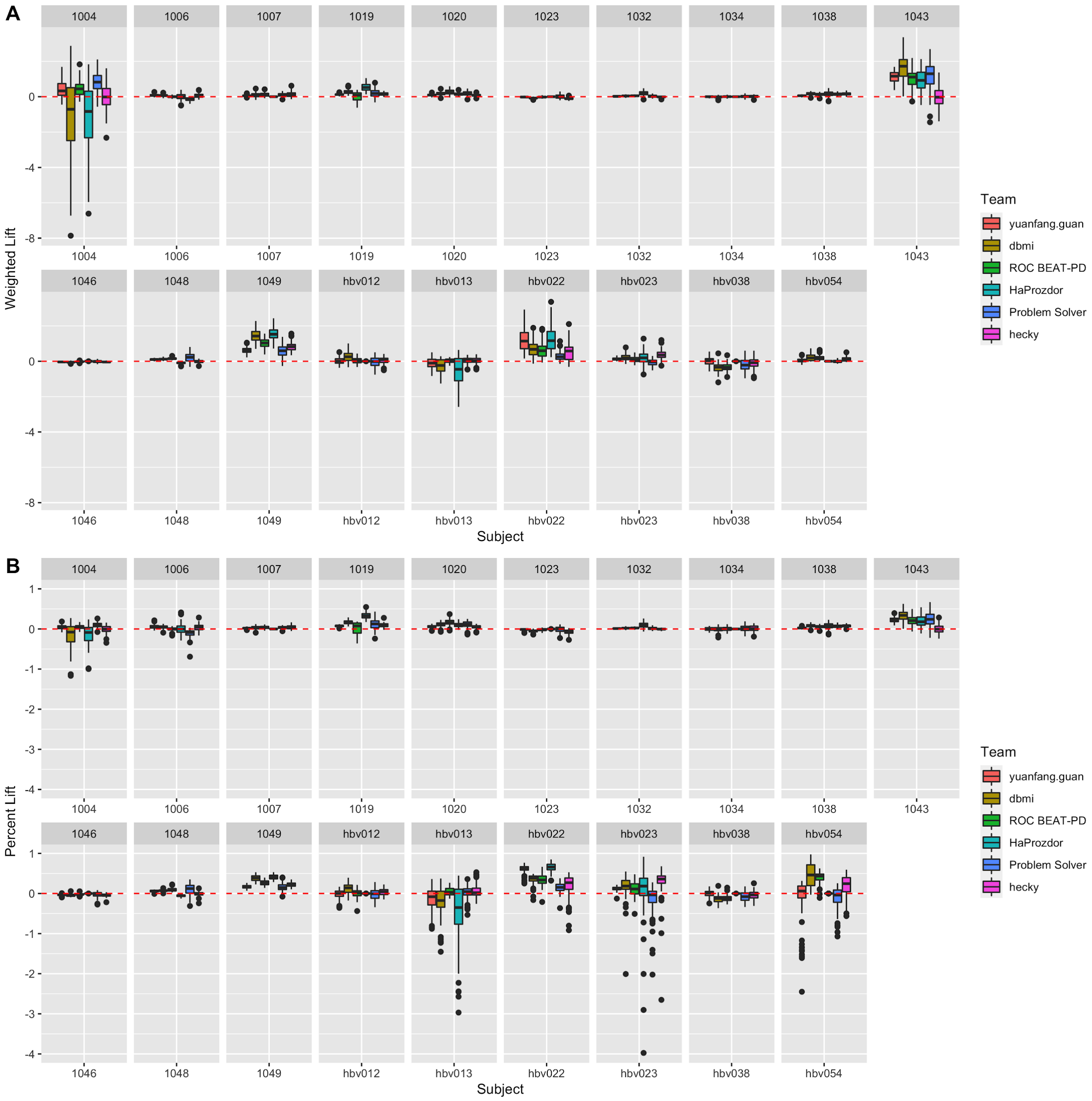


**Supplementary Figure 8**: Tremor subject-specific lift. (A) Weighted by sqrt(n) and (B) as a percentage of the null model MSE. Teams are ordered by overall rank.

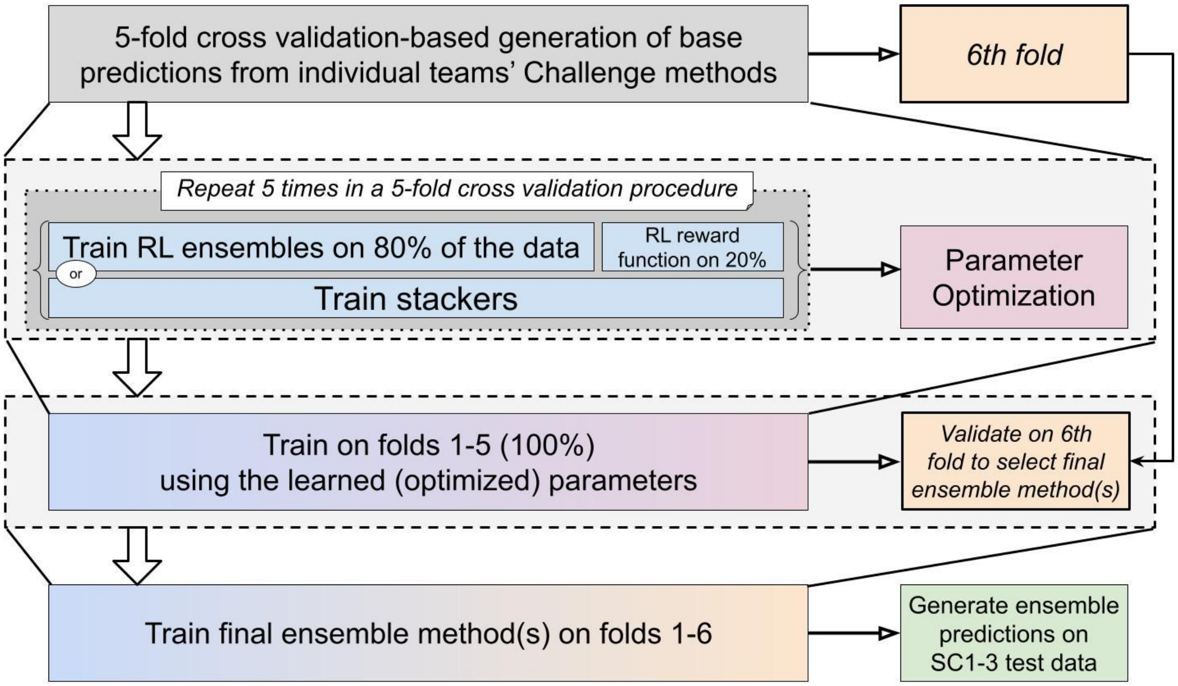


**Supplementary Figure 9**: Data-driven process used to train and evaluate heterogeneous ensembles for SC1-3.
